# Comparative Efficacy of Biologic Therapies for Plaque Psoriasis: A Network Meta-Analysis

**DOI:** 10.1101/2025.08.31.25334775

**Authors:** Huzaifa Saleem

## Abstract

**Background:** Biologics have significantly improved outcomes for patients with psoriasis. However, with multiple agents targeting different immunological pathways, comparative efficacy data is essential to guide optimal treatment selection.

**Objective:** To compare the efficacy of eight biologic therapies in achieving Psoriasis Area Severity Index 75 response in patients with plaque psoriasis.

**Methods:** A systematic review and network meta-analysis (NMA) was conducted per PRISMA-NMA guidelines. Open-access randomized controlled trials (RCTs) involving adults with plaque psoriasis treated with adalimumab, ustekinumab, secukinumab, ixekizumab, bimekizumab, guselkumab, risankizumab, or tildrakizumab were included. Psoriasis Area Severity Index 75 at approximately 12 weeks was the primary outcome. A frequentist random-effects model was used

**Results:** 29 RCTs (13923 participants) were included. 8 biologics were analyzed. All agents outperformed placebo. 11 of the combinations out of 325 had significant inconsistency. Transitivity assumptions based on randomization and study design were not addressed. Ixekizumab 80mg was the best treatment compared to placebo and all other biologics, while increasing dose of all drugs was not always associated with increasing efficacy except in the cases of Ustekinumab

## Introduction

Psoriasis is a chronic immune-mediated skin disorder that substantially impairs quality of life (1). Recent advances have introduced biologic therapies that target specific cytokines, including TNF-α, IL-12/23, IL-17, and IL-23. These therapies include; **TNF-**α **inhibitors**: Adalimumab**IL-12/23 inhibitors**: Ustekinumab, **IL-17 inhibitors**: Secukinumab, Ixekizumab, Bimekizumab, **IL-23 inhibitors**: Guselkumab, Risankizumab, Tildrakizumab

The variety of available treatments creates uncertainty for clinicians regarding optimal therapy. While direct head-to-head trials are limited, network meta-analysis (NMA) enables a comprehensive comparison by combining direct and indirect evidence. A meta-analysis published in 2022 ranked Infliximab as the best treatment in terms of achieving Psoriasis areas and severity index (PASI) 90%, followed by bimekizumab, ixekizumab and Risankizumab but was limited by a sample of only 19 RCTs (2). In a network meta-analysis published this year the efficacy of Janus kinase inhibitors in treating psoriatic arthritis was evaluated but was limited by a sample of only 5 studies (3). In another study evaluating the efficacy of biologics in Psoriasis Bimekizumab 320mg Q4W was the best treatment as compared to other biologics but again limited by sample of 19 Studies (4).

This study aims to compare and rank eight biologic agents for psoriasis based on their efficacy in achieving PASI 75.

## Methods

### Protocol and Registration

A protocol was developed after pilot literature search, but was not registered or published.

### Eligibility Criteria

- **Population**: Adults (>18 years) with moderate-to-severe plaque psoriasis. Only studies with both male and female participants were included.
- **Intervention/Comparators**: RCTs comparing adalimumab, ustekinumab, secukinumab, ixekizumab, bimekizumab, guselkumab, risankizumab, or tildrakizumab with placebo or each other.
- **Outcome**: PASI 75 (rang-75-100%) at approximately 12 weeks (range 8–16 weeks accepted).
- **Study Type**: Open-access RCTs only. No language restrictions applied.

### Information Sources and Search Strategy

Systematic search conducted in PubMed on March 25, 2025. Duplicates were removed using Rayyan. The following search terms were used:

“((((((((Guselkumab) AND (Psoriasis) AND (randomizedcontrolledtrial[Filter])) OR ((Ustekinumab) AND (Psoriasis) AND (randomizedcontrolledtrial[Filter]) AND (randomizedcontrolledtrial[Filter]))) OR ((Tildrakizumab) AND (Psoriasis) AND (randomizedcontrolledtrial[Filter]))) OR ((Secukinumab) AND (Psoriasis) AND (randomizedcontrolledtrial[Filter]))) OR ((Risankizumab) AND (Psoriasis) AND (randomizedcontrolledtrial[Filter]))) OR ((Ixekizumab) AND (Psoriasis) AND (randomizedcontrolledtrial[Filter]))) OR ((Bimekizumab) AND (Psoriasis) AND (randomizedcontrolledtrial[Filter]))) OR ((Adalimumab) AND (Psoriasis) AND (randomizedcontrolledtrial[Filter]))”

### Study Selection

Only Open access Randomized control trials were included, both head-to-head and placebo control trials were sought. Duplicates were removed using Rayyan, an automation tool. Only published studies were sought, no language limitation was set. Only open access studies were included. Full-text studies were screened using PICO criteria given above.

### Data Extraction

Extracted data included:

- Population demographics
- Intervention details (dose, mode, frequency)
- Outcome results (PASI 75-100%)
- Follow-up duration

### Data Synthesis and Statistical Analysis

A frequentist random-effects model was used to estimate Risk Ratios (RR) with 95% confidence intervals. Network plots illustrated geometry. League tables ranked treatments. Forest plots depicted treatment vs. placebo results. The size of the nodes in the network represents the number of studies that contribute to it. Same intervention with similar dose were combined in a single node. If a study reported results of multi-arm intervention, all were included in the network. METAINSIGHT was used to make and analyze the network meta-analysis.

### Assessment of Inconsistency and Transitivity

Local inconsistency was assessed using loop-specific approach. A p-value <0.05 indicated inconsistency. Transitivity assumption is evaluated using Clinical and methodological data as potential affect modifier across treatment comparison group. Clinical affect modifier in our study can be age, gender and dose of intervention, Methodological affect modifier can be bias in Randomization, if overall total number of patients do not have equal probability of being assigned to each intervention, then this can alter the effect size of the study. Transitivity argument was not addressed.

## Results

### Study Selection

303 articles were identified. After exclusions, 24 studies (5–28) were included (corresponding to 29 Randomized control trials). Study selection process is shown in **Figure 1: study selection process**. Characteristics of RCT are given in Supplementary file 1.

**Figure 1.**
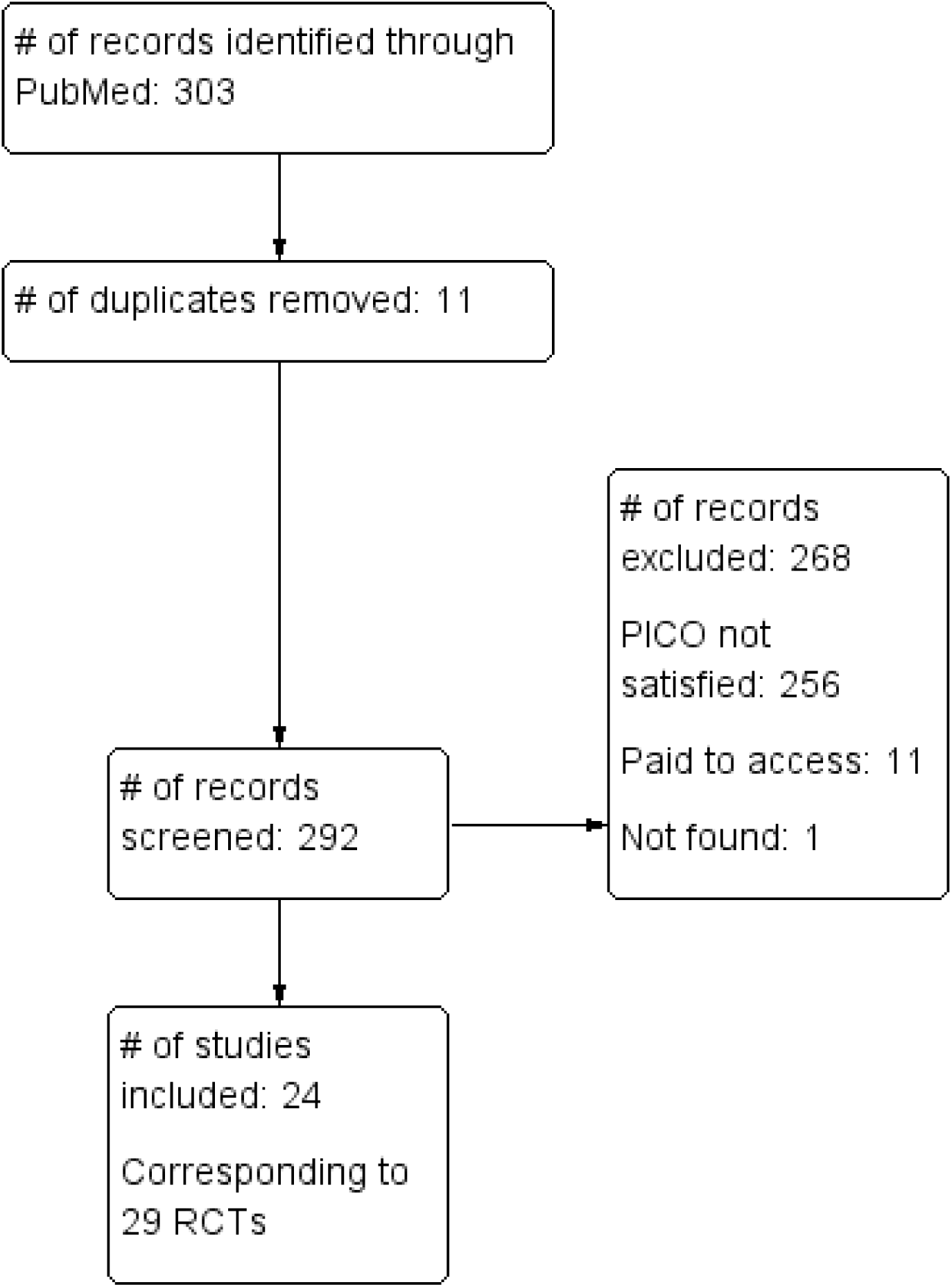
study selection process

### Study Characteristics

- A total of 29 RCTs were included (12 multi-arm studies)
- 19 head-to-head comparisons
- 18 placebo-controlled arms
- 13,923 participants (3218 placebo, 10710 active drug)
- Total # of new events in the network were 7194
- Mean treatment duration: weeks 12.6
- Supplementary file 2, shows forest plot of direct comparison from each trial.

### Network Geometry

The network included 8 interventions (26 when accounting for dosage difference). There were 325 Total possible pairwise comparison and 49 pairwise comparison with direct data. both direct and indirect comparison was present only for 27 pairs. Figure 2**: Network meta-analysis** shows the network structure.

**Figure 2.**
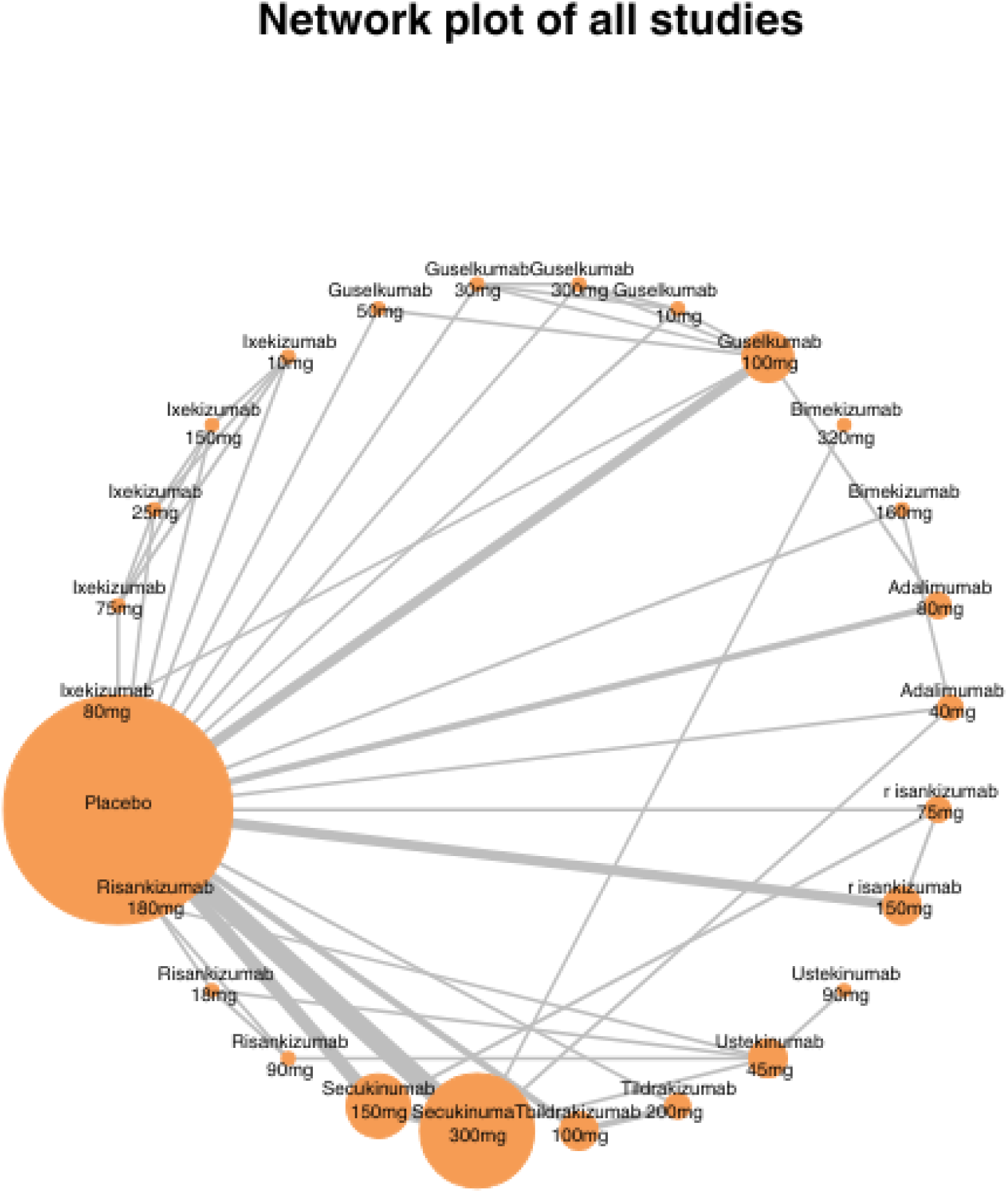
Network meta-analysis

### Relative Treatment Effects

All biologics were more effective than placebo based on pooled effect sizes of included studies see Figure 3**: Pooled efficacy for intervention vs placebo**:

**Figure 3.**
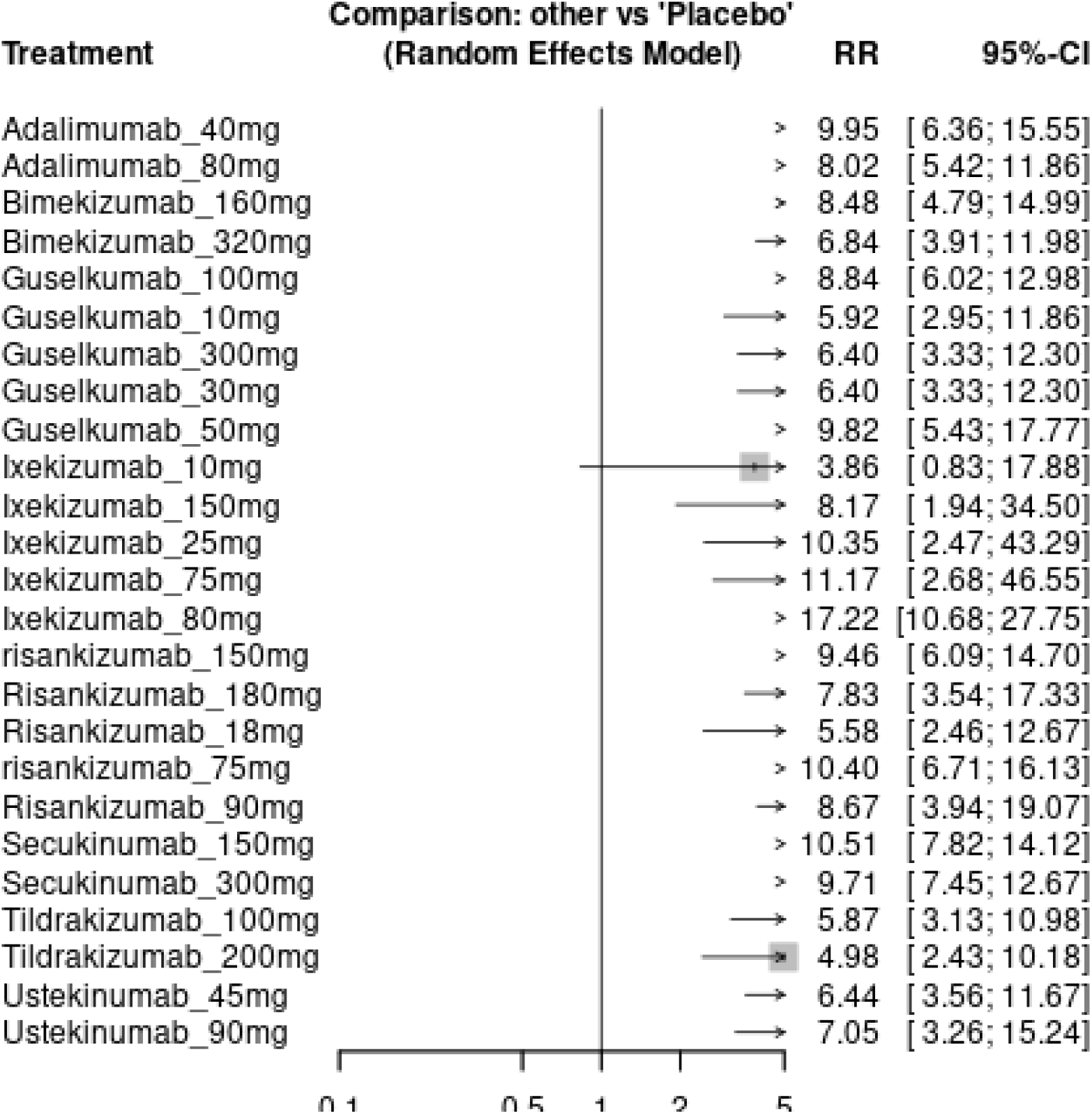
Pooled efficacy for intervention vs placebo

**Supplementary file 3**, shows a league table ranking each intervention according to their efficacy. League table uses direct and indirect evidence to compare each intervention. In the League Table, Data are RR (95% CI) in the column defining treatment compared with row defining treatment. Higher RR are desirable. RRs greater than 1 favors column defining the treatment. Ranking of the treatment follow order of the column, with column #1 being best and #26 being worst.

The order of best treatment for psoriasis is as follows: Ixekizumab 80mg, Secukinumab 150mg, Risankizumab 75mg, Adalimumab 40mg, Ixekizumab 75mg, Secukinumab 300mg, Guselkumab 50mg, Ixekizumab 25mg, Risankizumab 150mg, Guselkumab 100mg, Risankizumab 90mg, Bimekizumab 160mg, Ixekizumab 150mg, Adalimumab 80mg, Risankizumab 180mg, Ustekinumab 90mg, Bimekizumab 320mg, Guselkumab 300mg, Guselkumab 30mg, Ustekinumab 45mg, Tildrakizumab 100mg, Guselkumab 10mg, Risankzumab 18mg, Tildrakizumab 200mg, Ixekizumab 10mg, Pacebo.

### Inconsistency and Transitivity

There were a total 325 possible comparison but both direct and indirect comparison was present only for 27 pairs, there was inconsistency between direct and indirect evidence for 11 comparisons, see **Supplementary file 4: Inconsistency**. Clinical and methodological approach to the argument of Transitivity was not addressed.

## Discussion

This network meta-analysis (NMA) provides a comprehensive comparison of the short-term efficacy of eight biologic therapies in achieving PASI 75 response in patients with moderate-to-severe plaque psoriasis. The study included 29 randomized controlled trials (RCTs) involving 13923 participants making it one of the most extensive analyses of its kind to date. The findings demonstrate that all evaluated biologics—targeting TNF-α, IL-12/23, IL-17 and IL-23 pathways—were significantly more effective than placebo. Notably **ixekizumab 80mg** emerged as the most efficacious treatment, followed by **secukinumab 150mg** and **Risankizumab 75mg**. Rest of the ranking is given in Supplementary file 3.

The superior performance of ixekizumab aligns with its mechanism of action as an IL-17A inhibitor, a cytokine central to the pathogenesis of psoriasis. IL-17A plays a pivotal role in keratinocyte activation and the inflammatory cascade driving psoriatic plaques. The high efficacy of ixekizumab underscores the importance of targeting this pathway. Risankizumab an IL-23 inhibitor, also demonstrated strong efficacy, which is consistent with the critical role of IL-23 in sustaining Th17 cell responses. The ranking of these agents provides valuable insights for clinicians when selecting therapies for patients with plaque psoriasis.

Interestingly, the study revealed that increasing the dosage of most biologics did not consistently correlate with enhanced efficacy. Higher doses of ixekizumab (150mg) were less effective than lower doses (25mg). A trend also observed with risankizumab, bimekizumab, tildrakizumab, guselkumab, and adalimumab. The exception was Ustekinumab. This inconsistency suggests the presence of potential effect modifiers, such as variations in drug administration frequency, patient demographics, which may have influenced the results. Future research should explore these factors to optimize dosing strategies.

The study also highlighted significant inconsistency in 11 out of 325 pairwise comparisons, indicating variability between direct and indirect evidence. This inconsistency, along with the limited number of direct head-to-head comparisons, underscores the need for more robust clinical trials to validate these findings. Additionally, the transitivity assumption—critical for the validity of NMA—was not addressed. This may limit the generalizability of the results. Factors such as differences in study populations, outcome timing, and methodological quality may have introduced bias.

Several limitations of this analysis must be acknowledged. First, the variability in the timing of PASI 75 assessments across studies (8–16 weeks) may have affected outcome measurements. Second, the inclusion of only open-access RCTs and reliance on a single database (PubMed) may have introduced selection bias. Third, the lack of safety data precludes a comprehensive risk-benefit analysis. Critical appraisal is essential for clinical decision-making.

Despite these limitations, this NMA offers valuable insights into the relative efficacy of biologic therapies for plaque psoriasis. The findings support the use of ixekizumab and risankizumab as highly effective options, while also highlighting the need for further research to address inconsistencies and optimize dosing regimens. Future studies should incorporate long-term safety data, broader literature searches, and rigorous transitivity assessments to strengthen the evidence base.

## Conclusion

In conclusion, this study contributes to the growing body of evidence guiding treatment selection for plaque psoriasis. By ranking the efficacy of eight biologics, it aids clinicians in making informed decisions tailored to individual patient needs. However, the observed inconsistencies and limitations emphasize the importance of continued research to refine therapeutic strategies and improve patient outcomes.

## Data Availability

All data produced in the present work are contained in the manuscript

## Acknowledgement

None

## Conflict of interest

There is nothing to declare

## Clinical trial registration number

Not applicable, not a clinical trial

## Funding

None received or applied for

## Ethical approval and statement

Not applicable as no human or animal subjects were enrolled

## Ethical consideration

Not applicable

## Consent to publish

All data associated with this manuscript is owned by the author

---

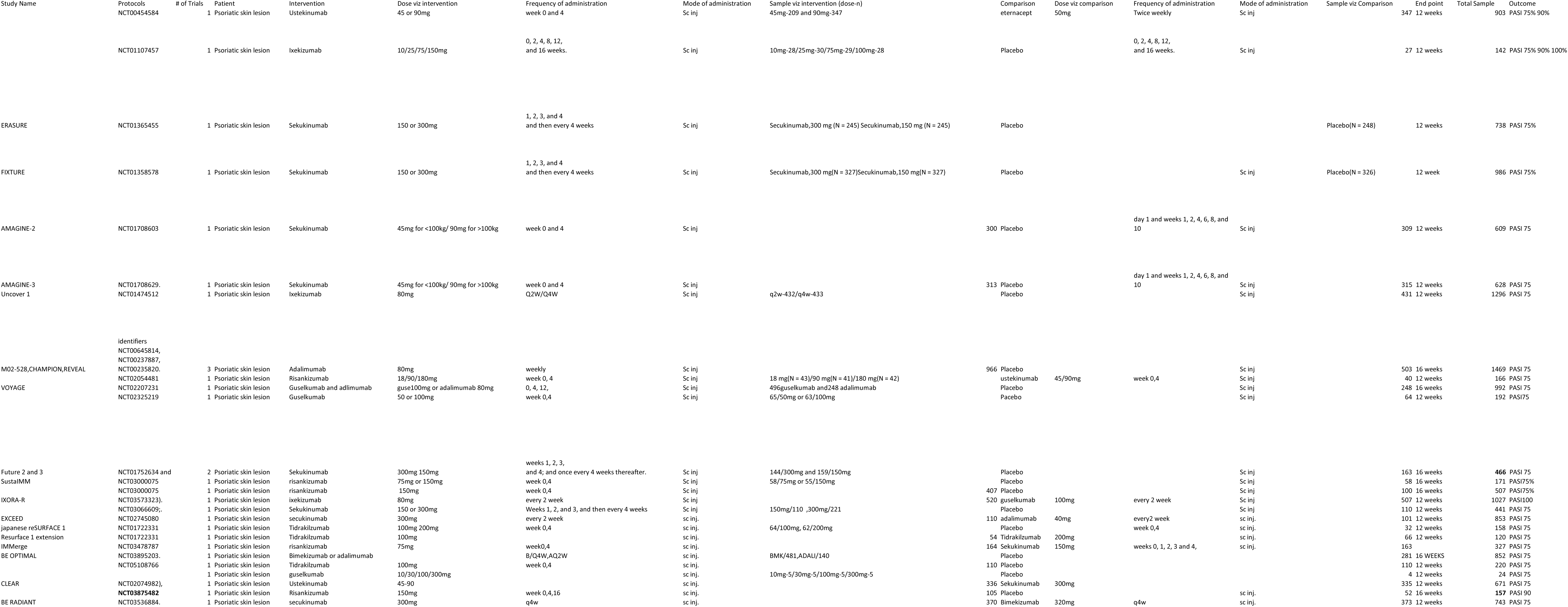

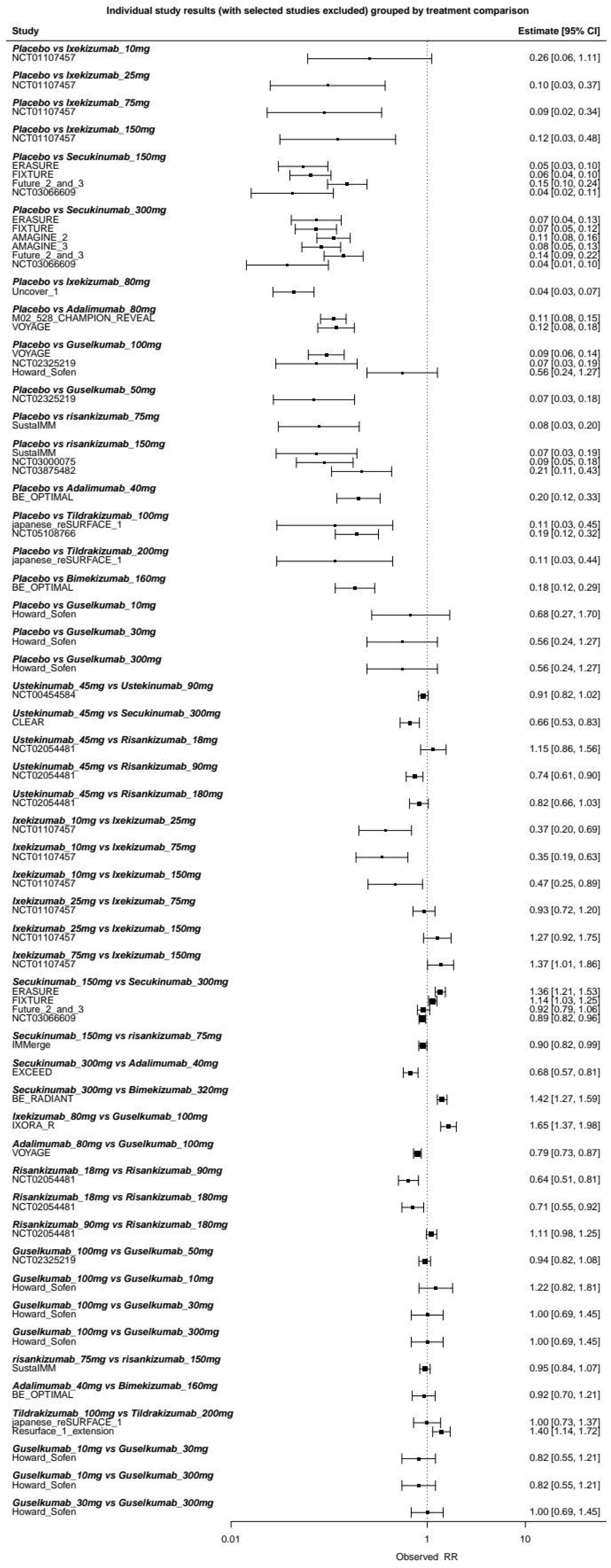

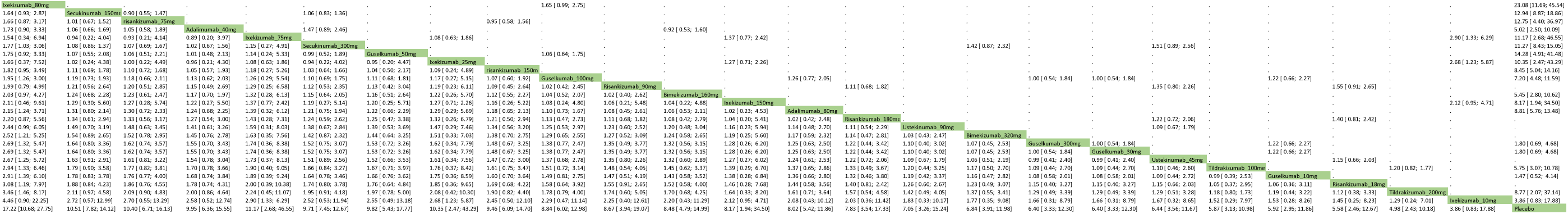

| Comparison | No.Studies | NMA | Direct | Indirect | Difference | 95CI_lower | 95CI_upper | pValue |
| --- | --- | --- | --- | --- | --- | --- | --- | --- |
| Adalimumab_40mg:Adalimumab_80mg | 0 | 0.215297 | NA | 0.215297 | NA | NA | NA | NA |
| Adalimumab_40mg:Bimekizumab_160mg | 1 | 0.159565 | -0.083077 | 1.94475 | -2.027827 | -3.619480948 | -0.436172911 | 0.012523 |
| Adalimumab_40mg:Bimekizumab_320mg | 0 | 0.374026 | NA | 0.374026 | NA | NA | NA | NA |
| Adalimumab_40mg:Guselkumab_100mg | 0 | 0.117959 | NA | 0.117959 | NA | NA | NA | NA |
| Adalimumab_40mg:Guselkumab_10mg | 0 | 0.519301 | NA | 0.519301 | NA | NA | NA | NA |
| Adalimumab_40mg:Guselkumab_300mg | 0 | 0.440335 | NA | 0.440335 | NA | NA | NA | NA |
| Adalimumab_40mg:Guselkumab_30mg | 0 | 0.440335 | NA | 0.440335 | NA | NA | NA | NA |
| Adalimumab_40mg:Guselkumab_50mg | 0 | 0.012414 | NA | 0.012414 | NA | NA | NA | NA |
| Adalimumab_40mg:Ixekizumab_10mg | 0 | 0.947162 | NA | 0.947162 | NA | NA | NA | NA |
| Adalimumab_40mg:Ixekizumab_150mg | 0 | 0.196491 | NA | 0.196491 | NA | NA | NA | NA |
| Adalimumab_40mg:Ixekizumab_25mg | 0 | -0.039898 | NA | -0.039898 | NA | NA | NA | NA |
| Adalimumab_40mg:Ixekizumab_75mg | 0 | -0.116359 | NA | -0.116359 | NA | NA | NA | NA |
| Adalimumab_40mg:Ixekizumab_80mg | 0 | -0.548891 | NA | -0.548891 | NA | NA | NA | NA |
| Adalimumab_40mg:Placebo | 1 | 2.297088 | 1.613003 | 2.770901 | -1.157898 | -2.066739427 | -0.249056637 | 0.012523 |
| Adalimumab_40mg:risankizumab_150mg | 0 | 0.050048 | NA | 0.050048 | NA | NA | NA | NA |
| Adalimumab_40mg:Risankizumab_180mg | 0 | 0.239039 | NA | 0.239039 | NA | NA | NA | NA |
| Adalimumab_40mg:Risankizumab_18mg | 0 | 0.577651 | NA | 0.577651 | NA | NA | NA | NA |
| Adalimumab_40mg:risankizumab_75mg | 0 | -0.044612 | NA | -0.044612 | NA | NA | NA | NA |
| Adalimumab_40mg:Risankizumab_90mg | 0 | 0.13698 | NA | 0.13698 | NA | NA | NA | NA |
| Adalimumab_40mg:Secukinumab_150mg | 0 | -0.055257 | NA | -0.055257 | NA | NA | NA | NA |
| Adalimumab_40mg:Secukinumab_300mg | 1 | 0.023665 | 0.388309 | -0.769589 | 1.157898 | 0.249056637 | 2.066739427 | 0.012523 |
| Adalimumab_40mg:Tildrakizumab_100mg | 0 | 0.527887 | NA | 0.527887 | NA | NA | NA | NA |
| Adalimumab_40mg:Tildrakizumab_200mg | 0 | 0.691978 | NA | 0.691978 | NA | NA | NA | NA |
| Adalimumab_40mg:Ustekinumab_45mg | 0 | 0.433871 | NA | 0.433871 | NA | NA | NA | NA |
| Adalimumab_40mg:Ustekinumab_90mg | 0 | 0.344444 | NA | 0.344444 | NA | NA | NA | NA |
| Adalimumab_80mg:Bimekizumab_160mg | 0 | -0.055732 | NA | -0.055732 | NA | NA | NA | NA |
| Adalimumab_80mg:Bimekizumab_320mg | 0 | 0.158729 | NA | 0.158729 | NA | NA | NA | NA |
| Adalimumab_80mg:Guselkumab_100mg | 1 | -0.097338 | -0.230178 | 0.247176 | -0.477353 | -1.403434135 | 0.448727256 | 0.312364 |
| Adalimumab_80mg:Guselkumab_10mg | 0 | 0.304004 | NA | 0.304004 | NA | NA | NA | NA |
| Adalimumab_80mg:Guselkumab_300mg | 0 | 0.225038 | NA | 0.225038 | NA | NA | NA | NA |
| Adalimumab_80mg:Guselkumab_30mg | 0 | 0.225038 | NA | 0.225038 | NA | NA | NA | NA |
| Adalimumab_80mg:Guselkumab_50mg | 0 | -0.202883 | NA | -0.202883 | NA | NA | NA | NA |
| Adalimumab_80mg:Ixekizumab_10mg | 0 | 0.731865 | NA | 0.731865 | NA | NA | NA | NA |
| Adalimumab_80mg:Ixekizumab_150mg | 0 | -0.018806 | NA | -0.018806 | NA | NA | NA | NA |
| Adalimumab_80mg:Ixekizumab_25mg | 0 | -0.255195 | NA | -0.255195 | NA | NA | NA | NA |
| Adalimumab_80mg:Ixekizumab_75mg | 0 | -0.331656 | NA | -0.331656 | NA | NA | NA | NA |
| Adalimumab_80mg:Ixekizumab_80mg | 0 | -0.764188 | NA | -0.764188 | NA | NA | NA | NA |
| Adalimumab_80mg:Placebo | 2 | 2.081791 | 2.176056 | 1.554831 | 0.621225 | -0.469669677 | 1.712118829 | 0.264367 |
| Adalimumab_80mg:risankizumab_150mg | 0 | -0.165249 | NA | -0.165249 | NA | NA | NA | NA |
| Adalimumab_80mg:Risankizumab_180mg | 0 | 0.023742 | NA | 0.023742 | NA | NA | NA | NA |
| Adalimumab_80mg:Risankizumab_18mg | 0 | 0.362354 | NA | 0.362354 | NA | NA | NA | NA |
| Adalimumab_80mg:risankizumab_75mg | 0 | -0.259909 | NA | -0.259909 | NA | NA | NA | NA |
| Adalimumab_80mg:Risankizumab_90mg | 0 | -0.078317 | NA | -0.078317 | NA | NA | NA | NA |
| Adalimumab_80mg:Secukinumab_150mg | 0 | -0.270554 | NA | -0.270554 | NA | NA | NA | NA |
| Adalimumab_80mg:Secukinumab_300mg | 0 | -0.191632 | NA | -0.191632 | NA | NA | NA | NA |
| Adalimumab_80mg:Tildrakizumab_100mg | 0 | 0.31259 | NA | 0.31259 | NA | NA | NA | NA |
| Adalimumab_80mg:Tildrakizumab_200mg | 0 | 0.476681 | NA | 0.476681 | NA | NA | NA | NA |
| Adalimumab_80mg:Ustekinumab_45mg | 0 | 0.218574 | NA | 0.218574 | NA | NA | NA | NA |
| Adalimumab_80mg:Ustekinumab_90mg | 0 | 0.129147 | NA | 0.129147 | NA | NA | NA | NA |
| Bimekizumab_160mg:Bimekizumab_320mg | 0 | 0.214462 | NA | 0.214462 | NA | NA | NA | NA |
| Bimekizumab_160mg:Guselkumab_100mg | 0 | -0.041606 | NA | -0.041606 | NA | NA | NA | NA |
| Bimekizumab_160mg:Guselkumab_10mg | 0 | 0.359736 | NA | 0.359736 | NA | NA | NA | NA |
| Bimekizumab_160mg:Guselkumab_300mg | 0 | 0.28077 | NA | 0.28077 | NA | NA | NA | NA |
| Bimekizumab_160mg:Guselkumab_30mg | 0 | 0.28077 | NA | 0.28077 | NA | NA | NA | NA |
| Bimekizumab_160mg:Guselkumab_50mg | 0 | -0.147151 | NA | -0.147151 | NA | NA | NA | NA |
| Bimekizumab_160mg:Ixekizumab_10mg | 0 | 0.787597 | NA | 0.787597 | NA | NA | NA | NA |
| Bimekizumab_160mg:Ixekizumab_150mg | 0 | 0.036926 | NA | 0.036926 | NA | NA | NA | NA |
| Bimekizumab_160mg:Ixekizumab_25mg | 0 | -0.199463 | NA | -0.199463 | NA | NA | NA | NA |
| Bimekizumab_160mg:Ixekizumab_75mg | 0 | -0.275924 | NA | -0.275924 | NA | NA | NA | NA |
| Bimekizumab_160mg:Ixekizumab_80mg | 0 | -0.708456 | NA | -0.708456 | NA | NA | NA | NA |
| Bimekizumab_160mg:Placebo | 1 | 2.137524 | 1.69608 | 3.332137 | -1.636058 | -2.920209507 | -0.351905784 | 0.012523 |
| Bimekizumab_160mg:risankizumab_150mg | 0 | -0.109516 | NA | -0.109516 | NA | NA | NA | NA |
| Bimekizumab_160mg:Risankizumab_180mg | 0 | 0.079475 | NA | 0.079475 | NA | NA | NA | NA |
| Bimekizumab_160mg:Risankizumab_18mg | 0 | 0.418086 | NA | 0.418086 | NA | NA | NA | NA |
| Bimekizumab_160mg:risankizumab_75mg | 0 | -0.204176 | NA | -0.204176 | NA | NA | NA | NA |
| Bimekizumab_160mg:Risankizumab_90mg | 0 | -0.022584 | NA | -0.022584 | NA | NA | NA | NA |
| Bimekizumab_160mg:Secukinumab_150mg | 0 | -0.214822 | NA | -0.214822 | NA | NA | NA | NA |
| Bimekizumab_160mg:Secukinumab_300mg | 0 | -0.1359 | NA | -0.1359 | NA | NA | NA | NA |
| Bimekizumab_160mg:Tildrakizumab_100mg | 0 | 0.368322 | NA | 0.368322 | NA | NA | NA | NA |
| Bimekizumab_160mg:Tildrakizumab_200mg | 0 | 0.532414 | NA | 0.532414 | NA | NA | NA | NA |
| Bimekizumab_160mg:Ustekinumab_45mg | 0 | 0.274307 | NA | 0.274307 | NA | NA | NA | NA |
| Bimekizumab_160mg:Ustekinumab_90mg | 0 | 0.18488 | NA | 0.18488 | NA | NA | NA | NA |
| Bimekizumab_320mg:Guselkumab_100mg | 0 | -0.256067 | NA | -0.256067 | NA | NA | NA | NA |
| Bimekizumab_320mg:Guselkumab_10mg | 0 | 0.145274 | NA | 0.145274 | NA | NA | NA | NA |
| Bimekizumab_320mg:Guselkumab_300mg | 0 | 0.066308 | NA | 0.066308 | NA | NA | NA | NA |
| Bimekizumab_320mg:Guselkumab_30mg | 0 | 0.066308 | NA | 0.066308 | NA | NA | NA | NA |
| Bimekizumab_320mg:Guselkumab_50mg | 0 | -0.361613 | NA | -0.361613 | NA | NA | NA | NA |
| Bimekizumab_320mg:Ixekizumab_10mg | 0 | 0.573135 | NA | 0.573135 | NA | NA | NA | NA |
| Bimekizumab_320mg:Ixekizumab_150mg | 0 | -0.177536 | NA | -0.177536 | NA | NA | NA | NA |
| Bimekizumab_320mg:Ixekizumab_25mg | 0 | -0.413925 | NA | -0.413925 | NA | NA | NA | NA |
| Bimekizumab_320mg:Ixekizumab_75mg | 0 | -0.490386 | NA | -0.490386 | NA | NA | NA | NA |
| Bimekizumab_320mg:Ixekizumab_80mg | 0 | -0.922918 | NA | -0.922918 | NA | NA | NA | NA |
| Bimekizumab_320mg:Placebo | 0 | 1.923062 | NA | 1.923062 | NA | NA | NA | NA |
| Bimekizumab_320mg:risankizumab_150mg | 0 | -0.323978 | NA | -0.323978 | NA | NA | NA | NA |
| Bimekizumab_320mg:Risankizumab_180mg | 0 | -0.134987 | NA | -0.134987 | NA | NA | NA | NA |
| Bimekizumab_320mg:Risankizumab_18mg | 0 | 0.203625 | NA | 0.203625 | NA | NA | NA | NA |
| Bimekizumab_320mg:risankizumab_75mg | 0 | -0.418638 | NA | -0.418638 | NA | NA | NA | NA |
| Bimekizumab_320mg:Risankizumab_90mg | 0 | -0.237046 | NA | -0.237046 | NA | NA | NA | NA |
| Bimekizumab_320mg:Secukinumab_150mg | 0 | -0.429284 | NA | -0.429284 | NA | NA | NA | NA |
| Bimekizumab_320mg:Secukinumab_300mg | 1 | -0.350362 | -0.350362 | NA | NA | NA | NA | NA |
| Bimekizumab_320mg:Tildrakizumab_100mg | 0 | 0.15386 | NA | 0.15386 | NA | NA | NA | NA |
| Bimekizumab_320mg:Tildrakizumab_200mg | 0 | 0.317952 | NA | 0.317952 | NA | NA | NA | NA |
| Bimekizumab_320mg:Ustekinumab_45mg | 0 | 0.059845 | NA | 0.059845 | NA | NA | NA | NA |
| Bimekizumab_320mg:Ustekinumab_90mg | 0 | -0.029582 | NA | -0.029582 | NA | NA | NA | NA |
| Guselkumab_100mg:Guselkumab_10mg | 1 | 0.401342 | 0.200671 | 6.556057 | -6.355386 | -9.849690912 | -2.861081603 | 0.000364 |
| Guselkumab_100mg:Guselkumab_300mg | 1 | 0.322376 | 0 | 3.796811 | -3.796811 | -5.88436591 | -1.709256788 | 0.000364 |
| Guselkumab_100mg:Guselkumab_30mg | 1 | 0.322376 | 0 | 3.796811 | -3.796811 | -5.88436591 | -1.709256788 | 0.000364 |
| Guselkumab_100mg:Guselkumab_50mg | 1 | -0.105545 | -0.058899 | -1.008414 | 0.949515 | -1.301654468 | 3.200684755 | 0.408414 |
| Guselkumab_100mg:Ixekizumab_10mg | 0 | 0.829203 | NA | 0.829203 | NA | NA | NA | NA |
| Guselkumab_100mg:Ixekizumab_150mg | 0 | 0.078532 | NA | 0.078532 | NA | NA | NA | NA |
| Guselkumab_100mg:Ixekizumab_25mg | 0 | -0.157857 | NA | -0.157857 | NA | NA | NA | NA |
| Guselkumab_100mg:Ixekizumab_75mg | 0 | -0.234318 | NA | -0.234318 | NA | NA | NA | NA |
| Guselkumab_100mg:Ixekizumab_80mg | 1 | -0.66685 | -0.499692 | -1.077129 | 0.577436 | -0.377326127 | 1.532198756 | 0.235868 |
| Guselkumab_100mg:Placebo | 3 | 2.179129 | 1.974186 | 2.564807 | -0.590621 | -1.39803145 | 0.21678907 | 0.151654 |
| Guselkumab_100mg:risankizumab_150mg | 0 | -0.067911 | NA | -0.067911 | NA | NA | NA | NA |
| Guselkumab_100mg:Risankizumab_180mg | 0 | 0.12108 | NA | 0.12108 | NA | NA | NA | NA |
| Guselkumab_100mg:Risankizumab_18mg | 0 | 0.459692 | NA | 0.459692 | NA | NA | NA | NA |
| Guselkumab_100mg:risankizumab_75mg | 0 | -0.162571 | NA | -0.162571 | NA | NA | NA | NA |
| Guselkumab_100mg:Risankizumab_90mg | 0 | 0.019021 | NA | 0.019021 | NA | NA | NA | NA |
| Guselkumab_100mg:Secukinumab_150mg | 0 | -0.173216 | NA | -0.173216 | NA | NA | NA | NA |
| Guselkumab_100mg:Secukinumab_300mg | 0 | -0.094294 | NA | -0.094294 | NA | NA | NA | NA |
| Guselkumab_100mg:Tildrakizumab_100mg | 0 | 0.409928 | NA | 0.409928 | NA | NA | NA | NA |
| Guselkumab_100mg:Tildrakizumab_200mg | 0 | 0.574019 | NA | 0.574019 | NA | NA | NA | NA |
| Guselkumab_100mg:Ustekinumab_45mg | 0 | 0.315912 | NA | 0.315912 | NA | NA | NA | NA |
| Guselkumab_100mg:Ustekinumab_90mg | 0 | 0.226485 | NA | 0.226485 | NA | NA | NA | NA |
| Guselkumab_10mg:Guselkumab_300mg | 1 | -0.078966 | -0.200671 | 10.27834 | -10.47901 | -16.24055907 | -4.717464252 | 0.000364 |
| Guselkumab_10mg:Guselkumab_30mg | 1 | -0.078966 | -0.200671 | 10.27834 | -10.47901 | -16.24055907 | -4.717464252 | 0.000364 |
| Guselkumab_10mg:Guselkumab_50mg | 0 | -0.506887 | NA | -0.506887 | NA | NA | NA | NA |
| Guselkumab_10mg:Ixekizumab_10mg | 0 | 0.427861 | NA | 0.427861 | NA | NA | NA | NA |
| Guselkumab_10mg:Ixekizumab_150mg | 0 | -0.32281 | NA | -0.32281 | NA | NA | NA | NA |
| Guselkumab_10mg:Ixekizumab_25mg | 0 | -0.559199 | NA | -0.559199 | NA | NA | NA | NA |
| Guselkumab_10mg:Ixekizumab_75mg | 0 | -0.63566 | NA | -0.63566 | NA | NA | NA | NA |
| Guselkumab_10mg:Ixekizumab_80mg | 0 | -1.068192 | NA | -1.068192 | NA | NA | NA | NA |
| Guselkumab_10mg:Placebo | 1 | 1.777787 | 0.387116 | 2.928656 | -2.54154 | -3.938923413 | -1.144155833 | 0.000364 |
| Guselkumab_10mg:risankizumab_150mg | 0 | -0.469253 | NA | -0.469253 | NA | NA | NA | NA |
| Guselkumab_10mg:Risankizumab_180mg | 0 | -0.280262 | NA | -0.280262 | NA | NA | NA | NA |
| Guselkumab_10mg:Risankizumab_18mg | 0 | 0.05835 | NA | 0.05835 | NA | NA | NA | NA |
| Guselkumab_10mg:risankizumab_75mg | 0 | -0.563913 | NA | -0.563913 | NA | NA | NA | NA |
| Guselkumab_10mg:Risankizumab_90mg | 0 | -0.382321 | NA | -0.382321 | NA | NA | NA | NA |
| Guselkumab_10mg:Secukinumab_150mg | 0 | -0.574558 | NA | -0.574558 | NA | NA | NA | NA |
| Guselkumab_10mg:Secukinumab_300mg | 0 | -0.495636 | NA | -0.495636 | NA | NA | NA | NA |
| Guselkumab_10mg:Tildrakizumab_100mg | 0 | 0.008586 | NA | 0.008586 | NA | NA | NA | NA |
| Guselkumab_10mg:Tildrakizumab_200mg | 0 | 0.172677 | NA | 0.172677 | NA | NA | NA | NA |
| Guselkumab_10mg:Ustekinumab_45mg | 0 | -0.08543 | NA | -0.08543 | NA | NA | NA | NA |
| Guselkumab_10mg:Ustekinumab_90mg | 0 | -0.174857 | NA | -0.174857 | NA | NA | NA | NA |
| Guselkumab_300mg:Guselkumab_30mg | 1 | 6.66E-16 | 0 | NA | NA | NA | NA | NA |
| Guselkumab_300mg:Guselkumab_50mg | 0 | -0.427921 | NA | -0.427921 | NA | NA | NA | NA |
| Guselkumab_300mg:Ixekizumab_10mg | 0 | 0.506827 | NA | 0.506827 | NA | NA | NA | NA |
| Guselkumab_300mg:Ixekizumab_150mg | 0 | -0.243844 | NA | -0.243844 | NA | NA | NA | NA |
| Guselkumab_300mg:Ixekizumab_25mg | 0 | -0.480233 | NA | -0.480233 | NA | NA | NA | NA |
| Guselkumab_300mg:Ixekizumab_75mg | 0 | -0.556694 | NA | -0.556694 | NA | NA | NA | NA |
| Guselkumab_300mg:Ixekizumab_80mg | 0 | -0.989226 | NA | -0.989226 | NA | NA | NA | NA |
| Guselkumab_300mg:Placebo | 1 | 1.856753 | 0.587787 | 2.968475 | -2.380688 | -3.6896325 | -1.071743242 | 0.000364 |
| Guselkumab_300mg:risankizumab_150mg | 0 | -0.390286 | NA | -0.390286 | NA | NA | NA | NA |
| Guselkumab_300mg:Risankizumab_180mg | 0 | -0.201295 | NA | -0.201295 | NA | NA | NA | NA |
| Guselkumab_300mg:Risankizumab_18mg | 0 | 0.137316 | NA | 0.137316 | NA | NA | NA | NA |
| Guselkumab_300mg:risankizumab_75mg | 0 | -0.484946 | NA | -0.484946 | NA | NA | NA | NA |
| Guselkumab_300mg:Risankizumab_90mg | 0 | -0.303354 | NA | -0.303354 | NA | NA | NA | NA |
| Guselkumab_300mg:Secukinumab_150mg | 0 | -0.495592 | NA | -0.495592 | NA | NA | NA | NA |
| Guselkumab_300mg:Secukinumab_300mg | 0 | -0.41667 | NA | -0.41667 | NA | NA | NA | NA |
| Guselkumab_300mg:Tildrakizumab_100mg | 0 | 0.087552 | NA | 0.087552 | NA | NA | NA | NA |
| Guselkumab_300mg:Tildrakizumab_200mg | 0 | 0.251644 | NA | 0.251644 | NA | NA | NA | NA |
| Guselkumab_300mg:Ustekinumab_45mg | 0 | -0.006463 | NA | -0.006463 | NA | NA | NA | NA |
| Guselkumab_300mg:Ustekinumab_90mg | 0 | -0.09589 | NA | -0.09589 | NA | NA | NA | NA |
| Guselkumab_30mg:Guselkumab_50mg | 0 | -0.427921 | NA | -0.427921 | NA | NA | NA | NA |
| Guselkumab_30mg:Ixekizumab_10mg | 0 | 0.506827 | NA | 0.506827 | NA | NA | NA | NA |
| Guselkumab_30mg:Ixekizumab_150mg | 0 | -0.243844 | NA | -0.243844 | NA | NA | NA | NA |
| Guselkumab_30mg:Ixekizumab_25mg | 0 | -0.480233 | NA | -0.480233 | NA | NA | NA | NA |
| Guselkumab_30mg:Ixekizumab_75mg | 0 | -0.556694 | NA | -0.556694 | NA | NA | NA | NA |
| Guselkumab_30mg:Ixekizumab_80mg | 0 | -0.989226 | NA | -0.989226 | NA | NA | NA | NA |
| Guselkumab_30mg:Placebo | 1 | 1.856753 | 0.587787 | 2.968475 | -2.380688 | -3.6896325 | -1.071743242 | 0.000364 |
| Guselkumab_30mg:risankizumab_150mg | 0 | -0.390286 | NA | -0.390286 | NA | NA | NA | NA |
| Guselkumab_30mg:Risankizumab_180mg | 0 | -0.201295 | NA | -0.201295 | NA | NA | NA | NA |
| Guselkumab_30mg:Risankizumab_18mg | 0 | 0.137316 | NA | 0.137316 | NA | NA | NA | NA |
| Guselkumab_30mg:risankizumab_75mg | 0 | -0.484946 | NA | -0.484946 | NA | NA | NA | NA |
| Guselkumab_30mg:Risankizumab_90mg | 0 | -0.303354 | NA | -0.303354 | NA | NA | NA | NA |
| Guselkumab_30mg:Secukinumab_150mg | 0 | -0.495592 | NA | -0.495592 | NA | NA | NA | NA |
| Guselkumab_30mg:Secukinumab_300mg | 0 | -0.41667 | NA | -0.41667 | NA | NA | NA | NA |
| Guselkumab_30mg:Tildrakizumab_100mg | 0 | 0.087552 | NA | 0.087552 | NA | NA | NA | NA |
| Guselkumab_30mg:Tildrakizumab_200mg | 0 | 0.251644 | NA | 0.251644 | NA | NA | NA | NA |
| Guselkumab_30mg:Ustekinumab_45mg | 0 | -0.006463 | NA | -0.006463 | NA | NA | NA | NA |
| Guselkumab_30mg:Ustekinumab_90mg | 0 | -0.09589 | NA | -0.09589 | NA | NA | NA | NA |
| Guselkumab_50mg:Ixekizumab_10mg | 0 | 0.934748 | NA | 0.934748 | NA | NA | NA | NA |
| Guselkumab_50mg:Ixekizumab_150mg | 0 | 0.184077 | NA | 0.184077 | NA | NA | NA | NA |
| Guselkumab_50mg:Ixekizumab_25mg | 0 | -0.052312 | NA | -0.052312 | NA | NA | NA | NA |
| Guselkumab_50mg:Ixekizumab_75mg | 0 | -0.128773 | NA | -0.128773 | NA | NA | NA | NA |
| Guselkumab_50mg:Ixekizumab_80mg | 0 | -0.561305 | NA | -0.561305 | NA | NA | NA | NA |
| Guselkumab_50mg:Placebo | 1 | 2.284674 | 2.658644 | 2.117371 | 0.541273 | -0.742011044 | 1.824557512 | 0.408414 |
| Guselkumab_50mg:risankizumab_150mg | 0 | 0.037635 | NA | 0.037635 | NA | NA | NA | NA |
| Guselkumab_50mg:Risankizumab_180mg | 0 | 0.226626 | NA | 0.226626 | NA | NA | NA | NA |
| Guselkumab_50mg:Risankizumab_18mg | 0 | 0.565237 | NA | 0.565237 | NA | NA | NA | NA |
| Guselkumab_50mg:risankizumab_75mg | 0 | -0.057025 | NA | -0.057025 | NA | NA | NA | NA |
| Guselkumab_50mg:Risankizumab_90mg | 0 | 0.124567 | NA | 0.124567 | NA | NA | NA | NA |
| Guselkumab_50mg:Secukinumab_150mg | 0 | -0.067671 | NA | -0.067671 | NA | NA | NA | NA |
| Guselkumab_50mg:Secukinumab_300mg | 0 | 0.011251 | NA | 0.011251 | NA | NA | NA | NA |
| Guselkumab_50mg:Tildrakizumab_100mg | 0 | 0.515473 | NA | 0.515473 | NA | NA | NA | NA |
| Guselkumab_50mg:Tildrakizumab_200mg | 0 | 0.679565 | NA | 0.679565 | NA | NA | NA | NA |
| Guselkumab_50mg:Ustekinumab_45mg | 0 | 0.421458 | NA | 0.421458 | NA | NA | NA | NA |
| Guselkumab_50mg:Ustekinumab_90mg | 0 | 0.332031 | NA | 0.332031 | NA | NA | NA | NA |
| Ixekizumab_10mg:Ixekizumab_150mg | 1 | -0.750671 | -0.750671 | NA | NA | NA | NA | NA |
| Ixekizumab_10mg:Ixekizumab_25mg | 1 | -0.98706 | -0.98706 | NA | NA | NA | NA | NA |
| Ixekizumab_10mg:Ixekizumab_75mg | 1 | -1.063521 | -1.063521 | NA | NA | NA | NA | NA |
| Ixekizumab_10mg:Ixekizumab_80mg | 0 | -1.496053 | NA | -1.496053 | NA | NA | NA | NA |
| Ixekizumab_10mg:Placebo | 1 | 1.349927 | 1.349927 | NA | NA | NA | NA | NA |
| Ixekizumab_10mg:risankizumab_150mg | 0 | -0.897113 | NA | -0.897113 | NA | NA | NA | NA |
| Ixekizumab_10mg:Risankizumab_180mg | 0 | -0.708122 | NA | -0.708122 | NA | NA | NA | NA |
| Ixekizumab_10mg:Risankizumab_18mg | 0 | -0.369511 | NA | -0.369511 | NA | NA | NA | NA |
| Ixekizumab_10mg:risankizumab_75mg | 0 | -0.991773 | NA | -0.991773 | NA | NA | NA | NA |
| Ixekizumab_10mg:Risankizumab_90mg | 0 | -0.810181 | NA | -0.810181 | NA | NA | NA | NA |
| Ixekizumab_10mg:Secukinumab_150mg | 0 | -1.002419 | NA | -1.002419 | NA | NA | NA | NA |
| Ixekizumab_10mg:Secukinumab_300mg | 0 | -0.923497 | NA | -0.923497 | NA | NA | NA | NA |
| Ixekizumab_10mg:Tildrakizumab_100mg | 0 | -0.419275 | NA | -0.419275 | NA | NA | NA | NA |
| Ixekizumab_10mg:Tildrakizumab_200mg | 0 | -0.255183 | NA | -0.255183 | NA | NA | NA | NA |
| Ixekizumab_10mg:Ustekinumab_45mg | 0 | -0.51329 | NA | -0.51329 | NA | NA | NA | NA |
| Ixekizumab_10mg:Ustekinumab_90mg | 0 | -0.602717 | NA | -0.602717 | NA | NA | NA | NA |
| Ixekizumab_150mg:Ixekizumab_25mg | 1 | -0.236389 | -0.236389 | NA | NA | NA | NA | NA |
| Ixekizumab_150mg:Ixekizumab_75mg | 1 | -0.31285 | -0.31285 | NA | NA | NA | NA | NA |
| Ixekizumab_150mg:Ixekizumab_80mg | 0 | -0.745382 | NA | -0.745382 | NA | NA | NA | NA |
| Ixekizumab_150mg:Placebo | 1 | 2.100598 | 2.100598 | NA | NA | NA | NA | NA |
| Ixekizumab_150mg:risankizumab_150mg | 0 | -0.146442 | NA | -0.146442 | NA | NA | NA | NA |
| Ixekizumab_150mg:Risankizumab_180mg | 0 | 0.042549 | NA | 0.042549 | NA | NA | NA | NA |
| Ixekizumab_150mg:Risankizumab_18mg | 0 | 0.38116 | NA | 0.38116 | NA | NA | NA | NA |
| Ixekizumab_150mg:risankizumab_75mg | 0 | -0.241102 | NA | -0.241102 | NA | NA | NA | NA |
| Ixekizumab_150mg:Risankizumab_90mg | 0 | -0.05951 | NA | -0.05951 | NA | NA | NA | NA |
| Ixekizumab_150mg:Secukinumab_150mg | 0 | -0.251748 | NA | -0.251748 | NA | NA | NA | NA |
| Ixekizumab_150mg:Secukinumab_300mg | 0 | -0.172826 | NA | -0.172826 | NA | NA | NA | NA |
| Ixekizumab_150mg:Tildrakizumab_100mg | 0 | 0.331396 | NA | 0.331396 | NA | NA | NA | NA |
| Ixekizumab_150mg:Tildrakizumab_200mg | 0 | 0.495488 | NA | 0.495488 | NA | NA | NA | NA |
| Ixekizumab_150mg:Ustekinumab_45mg | 0 | 0.237381 | NA | 0.237381 | NA | NA | NA | NA |
| Ixekizumab_150mg:Ustekinumab_90mg | 0 | 0.147954 | NA | 0.147954 | NA | NA | NA | NA |
| Ixekizumab_25mg:Ixekizumab_75mg | 1 | -0.076461 | -0.076461 | NA | NA | NA | NA | NA |
| Ixekizumab_25mg:Ixekizumab_80mg | 0 | -0.508993 | NA | -0.508993 | NA | NA | NA | NA |
| Ixekizumab_25mg:Placebo | 1 | 2.336987 | 2.336987 | NA | NA | NA | NA | NA |
| Ixekizumab_25mg:risankizumab_150mg | 0 | 0.089947 | NA | 0.089947 | NA | NA | NA | NA |
| Ixekizumab_25mg:Risankizumab_180mg | 0 | 0.278938 | NA | 0.278938 | NA | NA | NA | NA |
| Ixekizumab_25mg:Risankizumab_18mg | 0 | 0.617549 | NA | 0.617549 | NA | NA | NA | NA |
| Ixekizumab_25mg:risankizumab_75mg | 0 | -0.004713 | NA | -0.004713 | NA | NA | NA | NA |
| Ixekizumab_25mg:Risankizumab_90mg | 0 | 0.176879 | NA | 0.176879 | NA | NA | NA | NA |
| Ixekizumab_25mg:Secukinumab_150mg | 0 | -0.015359 | NA | -0.015359 | NA | NA | NA | NA |
| Ixekizumab_25mg:Secukinumab_300mg | 0 | 0.063563 | NA | 0.063563 | NA | NA | NA | NA |
| Ixekizumab_25mg:Tildrakizumab_100mg | 0 | 0.567785 | NA | 0.567785 | NA | NA | NA | NA |
| Ixekizumab_25mg:Tildrakizumab_200mg | 0 | 0.731877 | NA | 0.731877 | NA | NA | NA | NA |
| Ixekizumab_25mg:Ustekinumab_45mg | 0 | 0.47377 | NA | 0.47377 | NA | NA | NA | NA |
| Ixekizumab_25mg:Ustekinumab_90mg | 0 | 0.384343 | NA | 0.384343 | NA | NA | NA | NA |
| Ixekizumab_75mg:Ixekizumab_80mg | 0 | -0.432532 | NA | -0.432532 | NA | NA | NA | NA |
| Ixekizumab_75mg:Placebo | 1 | 2.413448 | 2.413448 | NA | NA | NA | NA | NA |
| Ixekizumab_75mg:risankizumab_150mg | 0 | 0.166408 | NA | 0.166408 | NA | NA | NA | NA |
| Ixekizumab_75mg:Risankizumab_180mg | 0 | 0.355399 | NA | 0.355399 | NA | NA | NA | NA |
| Ixekizumab_75mg:Risankizumab_18mg | 0 | 0.69401 | NA | 0.69401 | NA | NA | NA | NA |
| Ixekizumab_75mg:risankizumab_75mg | 0 | 0.071748 | NA | 0.071748 | NA | NA | NA | NA |
| Ixekizumab_75mg:Risankizumab_90mg | 0 | 0.25334 | NA | 0.25334 | NA | NA | NA | NA |
| Ixekizumab_75mg:Secukinumab_150mg | 0 | 0.061102 | NA | 0.061102 | NA | NA | NA | NA |
| Ixekizumab_75mg:Secukinumab_300mg | 0 | 0.140024 | NA | 0.140024 | NA | NA | NA | NA |
| Ixekizumab_75mg:Tildrakizumab_100mg | 0 | 0.644246 | NA | 0.644246 | NA | NA | NA | NA |
| Ixekizumab_75mg:Tildrakizumab_200mg | 0 | 0.808338 | NA | 0.808338 | NA | NA | NA | NA |
| Ixekizumab_75mg:Ustekinumab_45mg | 0 | 0.550231 | NA | 0.550231 | NA | NA | NA | NA |
| Ixekizumab_75mg:Ustekinumab_90mg | 0 | 0.460804 | NA | 0.460804 | NA | NA | NA | NA |
| Ixekizumab_80mg:Placebo | 1 | 2.845979 | 3.13879 | 2.561354 | 0.577436 | -0.377326127 | 1.532198756 | 0.235868 |
| Ixekizumab_80mg:risankizumab_150mg | 0 | 0.59894 | NA | 0.59894 | NA | NA | NA | NA |
| Ixekizumab_80mg:Risankizumab_180mg | 0 | 0.787931 | NA | 0.787931 | NA | NA | NA | NA |
| Ixekizumab_80mg:Risankizumab_18mg | 0 | 1.126542 | NA | 1.126542 | NA | NA | NA | NA |
| Ixekizumab_80mg:risankizumab_75mg | 0 | 0.50428 | NA | 0.50428 | NA | NA | NA | NA |
| Ixekizumab_80mg:Risankizumab_90mg | 0 | 0.685872 | NA | 0.685872 | NA | NA | NA | NA |
| Ixekizumab_80mg:Secukinumab_150mg | 0 | 0.493634 | NA | 0.493634 | NA | NA | NA | NA |
| Ixekizumab_80mg:Secukinumab_300mg | 0 | 0.572556 | NA | 0.572556 | NA | NA | NA | NA |
| Ixekizumab_80mg:Tildrakizumab_100mg | 0 | 1.076778 | NA | 1.076778 | NA | NA | NA | NA |
| Ixekizumab_80mg:Tildrakizumab_200mg | 0 | 1.240869 | NA | 1.240869 | NA | NA | NA | NA |
| Ixekizumab_80mg:Ustekinumab_45mg | 0 | 0.982763 | NA | 0.982763 | NA | NA | NA | NA |
| Ixekizumab_80mg:Ustekinumab_90mg | 0 | 0.893336 | NA | 0.893336 | NA | NA | NA | NA |
| risankizumab_150mg:Placebo | 3 | 2.24704 | 2.133753 | 2.551032 | -0.417279 | -1.408882037 | 0.574323226 | 0.409497 |
| Risankizumab_180mg:Placebo | 0 | 2.058049 | NA | 2.058049 | NA | NA | NA | NA |
| Risankizumab_18mg:Placebo | 0 | 1.719437 | NA | 1.719437 | NA | NA | NA | NA |
| risankizumab_75mg:Placebo | 1 | 2.3417 | 2.545531 | 2.299982 | 0.245549 | -0.923018076 | 1.414115752 | 0.680454 |
| Risankizumab_90mg:Placebo | 0 | 2.160108 | NA | 2.160108 | NA | NA | NA | NA |
| Secukinumab_150mg:Placebo | 4 | 2.352346 | 2.560105 | 2.021453 | 0.538652 | -0.068553649 | 1.145858626 | 0.08209 |
| Secukinumab_300mg:Placebo | 6 | 2.273423 | 2.421758 | 1.489792 | 0.931966 | 0.20599244 | 1.657939645 | 0.011866 |
| Tildrakizumab_100mg:Placebo | 2 | 1.769202 | 1.749661 | 6.421032 | -4.671371 | -14.38823545 | 5.04549296 | 0.346064 |
| Tildrakizumab_200mg:Placebo | 1 | 1.60511 | 2.171815 | 1.420393 | 0.751422 | -0.910105052 | 2.412949167 | 0.375408 |
| Ustekinumab_45mg:Placebo | 0 | 1.863217 | NA | 1.863217 | NA | NA | NA | NA |
| Ustekinumab_90mg:Placebo | 0 | 1.952644 | NA | 1.952644 | NA | NA | NA | NA |
| risankizumab_150mg:Risankizumab_180mg | 0 | 0.188991 | NA | 0.188991 | NA | NA | NA | NA |
| risankizumab_150mg:Risankizumab_18mg | 0 | 0.527603 | NA | 0.527603 | NA | NA | NA | NA |
| risankizumab_150mg:risankizumab_75mg | 1 | -0.09466 | 0.05311 | -0.510381 | 0.563491 | -0.402870777 | 1.529852104 | 0.253094 |
| risankizumab_150mg:Risankizumab_90mg | 0 | 0.086932 | NA | 0.086932 | NA | NA | NA | NA |
| risankizumab_150mg:Secukinumab_150mg | 0 | -0.105306 | NA | -0.105306 | NA | NA | NA | NA |
| risankizumab_150mg:Secukinumab_300mg | 0 | -0.026384 | NA | -0.026384 | NA | NA | NA | NA |
| risankizumab_150mg:Tildrakizumab_100mg | 0 | 0.477838 | NA | 0.477838 | NA | NA | NA | NA |
| risankizumab_150mg:Tildrakizumab_200mg | 0 | 0.64193 | NA | 0.64193 | NA | NA | NA | NA |
| risankizumab_150mg:Ustekinumab_45mg | 0 | 0.383823 | NA | 0.383823 | NA | NA | NA | NA |
| risankizumab_150mg:Ustekinumab_90mg | 0 | 0.294396 | NA | 0.294396 | NA | NA | NA | NA |
| Risankizumab_180mg:Risankizumab_18mg | 1 | 0.338612 | 0.338612 | NA | NA | NA | NA | NA |
| Risankizumab_180mg:risankizumab_75mg | 0 | -0.283651 | NA | -0.283651 | NA | NA | NA | NA |
| Risankizumab_180mg:Risankizumab_90mg | 1 | -0.102059 | -0.102059 | NA | NA | NA | NA | NA |
| Risankizumab_180mg:Secukinumab_150mg | 0 | -0.294297 | NA | -0.294297 | NA | NA | NA | NA |
| Risankizumab_180mg:Secukinumab_300mg | 0 | -0.215375 | NA | -0.215375 | NA | NA | NA | NA |
| Risankizumab_180mg:Tildrakizumab_100mg | 0 | 0.288847 | NA | 0.288847 | NA | NA | NA | NA |
| Risankizumab_180mg:Tildrakizumab_200mg | 0 | 0.452939 | NA | 0.452939 | NA | NA | NA | NA |
| Risankizumab_180mg:Ustekinumab_45mg | 1 | 0.194832 | 0.194832 | NA | NA | NA | NA | NA |
| Risankizumab_180mg:Ustekinumab_90mg | 0 | 0.105405 | NA | 0.105405 | NA | NA | NA | NA |
| Risankizumab_18mg:risankizumab_75mg | 0 | -0.622263 | NA | -0.622263 | NA | NA | NA | NA |
| Risankizumab_18mg:Risankizumab_90mg | 1 | -0.440671 | -0.440671 | NA | NA | NA | NA | NA |
| Risankizumab_18mg:Secukinumab_150mg | 0 | -0.632908 | NA | -0.632908 | NA | NA | NA | NA |
| Risankizumab_18mg:Secukinumab_300mg | 0 | -0.553986 | NA | -0.553986 | NA | NA | NA | NA |
| Risankizumab_18mg:Tildrakizumab_100mg | 0 | -0.049764 | NA | -0.049764 | NA | NA | NA | NA |
| Risankizumab_18mg:Tildrakizumab_200mg | 0 | 0.114327 | NA | 0.114327 | NA | NA | NA | NA |
| Risankizumab_18mg:Ustekinumab_45mg | 1 | -0.14378 | -0.14378 | NA | NA | NA | NA | NA |
| Risankizumab_18mg:Ustekinumab_90mg | 0 | -0.233207 | NA | -0.233207 | NA | NA | NA | NA |
| risankizumab_75mg:Risankizumab_90mg | 0 | 0.181592 | NA | 0.181592 | NA | NA | NA | NA |
| risankizumab_75mg:Secukinumab_150mg | 1 | -0.010646 | 0.103083 | -0.269644 | 0.372727 | -0.513004033 | 1.258458885 | 0.409497 |
| risankizumab_75mg:Secukinumab_300mg | 0 | 0.068276 | NA | 0.068276 | NA | NA | NA | NA |
| risankizumab_75mg:Tildrakizumab_100mg | 0 | 0.572498 | NA | 0.572498 | NA | NA | NA | NA |
| risankizumab_75mg:Tildrakizumab_200mg | 0 | 0.73659 | NA | 0.73659 | NA | NA | NA | NA |
| risankizumab_75mg:Ustekinumab_45mg | 0 | 0.478483 | NA | 0.478483 | NA | NA | NA | NA |
| risankizumab_75mg:Ustekinumab_90mg | 0 | 0.389056 | NA | 0.389056 | NA | NA | NA | NA |
| Risankizumab_90mg:Secukinumab_150mg | 0 | -0.192238 | NA | -0.192238 | NA | NA | NA | NA |
| Risankizumab_90mg:Secukinumab_300mg | 0 | -0.113316 | NA | -0.113316 | NA | NA | NA | NA |
| Risankizumab_90mg:Tildrakizumab_100mg | 0 | 0.390906 | NA | 0.390906 | NA | NA | NA | NA |
| Risankizumab_90mg:Tildrakizumab_200mg | 0 | 0.554998 | NA | 0.554998 | NA | NA | NA | NA |
| Risankizumab_90mg:Ustekinumab_45mg | 1 | 0.296891 | 0.296891 | NA | NA | NA | NA | NA |
| Risankizumab_90mg:Ustekinumab_90mg | 0 | 0.207464 | NA | 0.207464 | NA | NA | NA | NA |
| Secukinumab_150mg:Secukinumab_300mg | 4 | 0.078922 | 0.05869 | 0.292124 | -0.233433 | -1.070325986 | 0.603459372 | 0.584592 |
| Secukinumab_150mg:Tildrakizumab_100mg | 0 | 0.583144 | NA | 0.583144 | NA | NA | NA | NA |
| Secukinumab_150mg:Tildrakizumab_200mg | 0 | 0.747236 | NA | 0.747236 | NA | NA | NA | NA |
| Secukinumab_150mg:Ustekinumab_45mg | 0 | 0.489129 | NA | 0.489129 | NA | NA | NA | NA |
| Secukinumab_150mg:Ustekinumab_90mg | 0 | 0.399702 | NA | 0.399702 | NA | NA | NA | NA |
| Secukinumab_300mg:Tildrakizumab_100mg | 0 | 0.504222 | NA | 0.504222 | NA | NA | NA | NA |
| Secukinumab_300mg:Tildrakizumab_200mg | 0 | 0.668313 | NA | 0.668313 | NA | NA | NA | NA |
| Secukinumab_300mg:Ustekinumab_45mg | 1 | 0.410207 | 0.410207 | NA | NA | NA | NA | NA |
| Secukinumab_300mg:Ustekinumab_90mg | 0 | 0.320779 | NA | 0.320779 | NA | NA | NA | NA |
| Tildrakizumab_100mg:Tildrakizumab_200mg | 2 | 0.164092 | 0.18208 | -1.04212 | 1.2242 | -1.965152256 | 4.413552678 | 0.451863 |
| Tildrakizumab_100mg:Ustekinumab_45mg | 0 | -0.094015 | NA | -0.094015 | NA | NA | NA | NA |
| Tildrakizumab_100mg:Ustekinumab_90mg | 0 | -0.183442 | NA | -0.183442 | NA | NA | NA | NA |
| Tildrakizumab_200mg:Ustekinumab_45mg | 0 | -0.258107 | NA | -0.258107 | NA | NA | NA | NA |
| Tildrakizumab_200mg:Ustekinumab_90mg | 0 | -0.347534 | NA | -0.347534 | NA | NA | NA | NA |
| Ustekinumab_45mg:Ustekinumab_90mg | 1 | -0.089427 | -0.089427 | NA | NA | NA | NA | NA |

## Notes

### Competing Interest Statement

The authors have declared no competing interest.

### Funding Statement

This study did not receive any funding

